# Vitamin D receptor (VDR) gene *Fok*I, *Bsm*I, *Apa*I and *Taq*I polymorphisms and osteoporosis risk: a meta-analysis

**DOI:** 10.1101/19009746

**Authors:** Upendra Yadav, Pradeep Kumar, Vandana Rai

## Abstract

Osteoporosis is a bone disease characterized by low bone density. The prevalence of osteoporosis varies between different populations and ethnic groups. Numerous studies have investigated the relationship between VDR gene polymorphisms and osteoporosis across ethnic populations. Present meta-analysis aims to comprehensively evaluate the influence of common *Fok*I, *Bsm*I, *Apa*I and *Taq*I VDR gene polymorphisms and osteoporosis. PubMed, Google Scholar, Springer Link and Elsevier databases were searched for eligible studies and all statistical calculations were performed by Open Meta-Analyst software. Studies investigated *Bsm*I (65 studies; 6,880 case/ 8,049 control), *Apa*I (31 studies; 3,763 case/ 3,934 control), *Fok*I (18 studies; 1,895 case/ 1,722 control), and *Taq*I (26 studies; 2,458 case/ 2,895 control) polymorphisms were included in the present meta-analysis. Results of meta-analysis revealed significant association between dominant model of *Fok*I (OR_ff+Ff vs. FF_= 1.19, 95% CI= 1.04-1.36, p= 0.01, I^2^= 39.36%) in overall analysis and recessive model of Caucasian population of *Taq*I polymorphism (OR_TT+Tt vs. tt_= 1.35, 95% CI= 1.11-1.63, p= 0.002, I^2^= 50.07%). While no such effect is found in any other genetic model in any other gene polymorphisms of the overall analyses or sub-group analyses. In conclusion, we found the *Fok*I polymorphism is associated with osteoporosis in overall analysis, also the *Taq*I polymorphism is a risk factor for the Caucasian population.

## Introduction

Bone is a very active tissue that maintain itself by continuously formation and reabsorption. To maintain this equilibrium osteoclasts, osteoblasts and osteocytes plays important role by performing bone resorption, formation and maintenance respectively [1]. Osteoporosis is a bone disease characterized by low bone density caused by increased activity of osteoclasts and decreased bone turnover [2]. The prevalence of osteoporosis greatly varies in different populations and ethnic groups [3]. Age and gender are the major contributing factor in the occurrence of osteoporosis. Worldwide every one women out of three over age of 50 will experience osteoporotic fractures in comparison to one in five men of the same age group [4]. The interaction between genetic and environmental factors are believed to play a central role in the etiology of osteoporosis [5, 6]. Among environmental factors exercise and calcium intake are crucial risk factor for the development of osteoporosis [5]. Now several evidences very well established that genetic factors play important role in the development of osteoporosis like- (i) female offspring of osteoporotic women have lower bone density in comparison with that of those with normal bone density values [7], (ii) male offspring of men who are diagnosed with idiopathic osteoporosis have lower bone mineral density (BMD) in comparison with that of men with normal bone density values [8] and (iii) studies of female twins have shown heritability of bone mineral density (BMD) to be 57% to 92% [9, 10].

Published studies have reported a list of effective genes for osteoporosis; the most important of which are vitamin D receptor gene (VDR), estrogen receptor alpha (ESRα), interleukin -6 (IL-6), Collagen type I (COLIA1), LDL receptor-related protein 5 (LRP5) [11, 12] etc. *VDR* gene polymorphisms have been reported to associated with the development of several bone diseases, multiple sclerosis, vitamin D dependent rickets type II and other complex diseases [13]. However, the mechanism by which the *VDR* gene influences bone mass has not been fully elucidated.

In humans, *VDR* gene is mapped at chromosome 12 (12q12-q14), it has 11 exons and spans ∼75 kb of genomic DNA. The most studied *VDR* gene polymorphisms are-*Bsm*I (rs1544410), *Apa*I (rs7975232), *Fok*I (rs10735810), and *Taq*I (rs731236).Although several studies between osteoporosis and *VDR* gene polymorphisms have been published, the results have been contradictory [14, 15], possibly because of variations in study design, small sample sizes, varying ethnic backgrounds, or environmental factors. So, we performed a meta-analysis to elucidate the role of these genetic polymorphisms in the etiology of osteoporosis.

## Materials and methods

Different databases (PubMed; Google Scholar, SpringerLink) were searched up to December 31, 2018 with the keywords ‘vitamin D receptor gene’, ‘*Bsm*I’, ‘*Apa*I’, ‘*Fok*I’, ‘*Taq*I’, ‘VDR’, and ‘osteoporosis’. The retrieved studies were published from 1995 to 2018 and we thoroughly examined all retrieved articles to assess their appropriateness for inclusion in the present meta-analysis.

### Inclusion and exclusion criteria

Eligible studies had to meet all of the following criteria: (a) the study should be a case-control study and (b) the articles must report the sample size, distribution of genotypes. The following exclusion criteria were used for excluding studies: (a) studies conducted on the animal model system; (b) studies that contained duplicate data; (c) no usable data reported; (d) only cases were reported; and (e) book chapters or reviews articles.

### Data extraction

The following information were extracted from all the selected articles: (a) the name of the first author; (b) year of publication; (c) country of study; (d) source of the control group; and (e) distribution of genotypes in case and control groups. We also evaluated whether the genotype distributions of control population of all the included studies were in agreement of Hardy–Weinberg equilibrium (HWE).

### Statistical analysis

Meta-analysis was done according to the method given in Rai et al. [16]. Pooled odds ratio (OR) with its corresponding 95% confidence interval (CI) to investigate the association between different *VDR* gene polymorphisms and risk of osteoporosis. Heterogeneity, Publication bias and subgroup analysis were done as per the method given in the Rai et al. [16].

## Results

For meta-analysis, we followed PRISMA guidelines. Figure 1 presents a flow chart of the retrieved studies and the studies included and excluded, with specifying reasons in the meta-analysis (Figure 1). After applying inclusion and exclusion criteria, 81 studies were found suitable for the inclusion in the present meta-analysis. Out of 81 included studies, *Bsm*I, *Apa*I, *Fok*I and *Taq*I polymorphism were investigated in 65, 31, 18 and 26 studies respectively.

**Figure 1.**
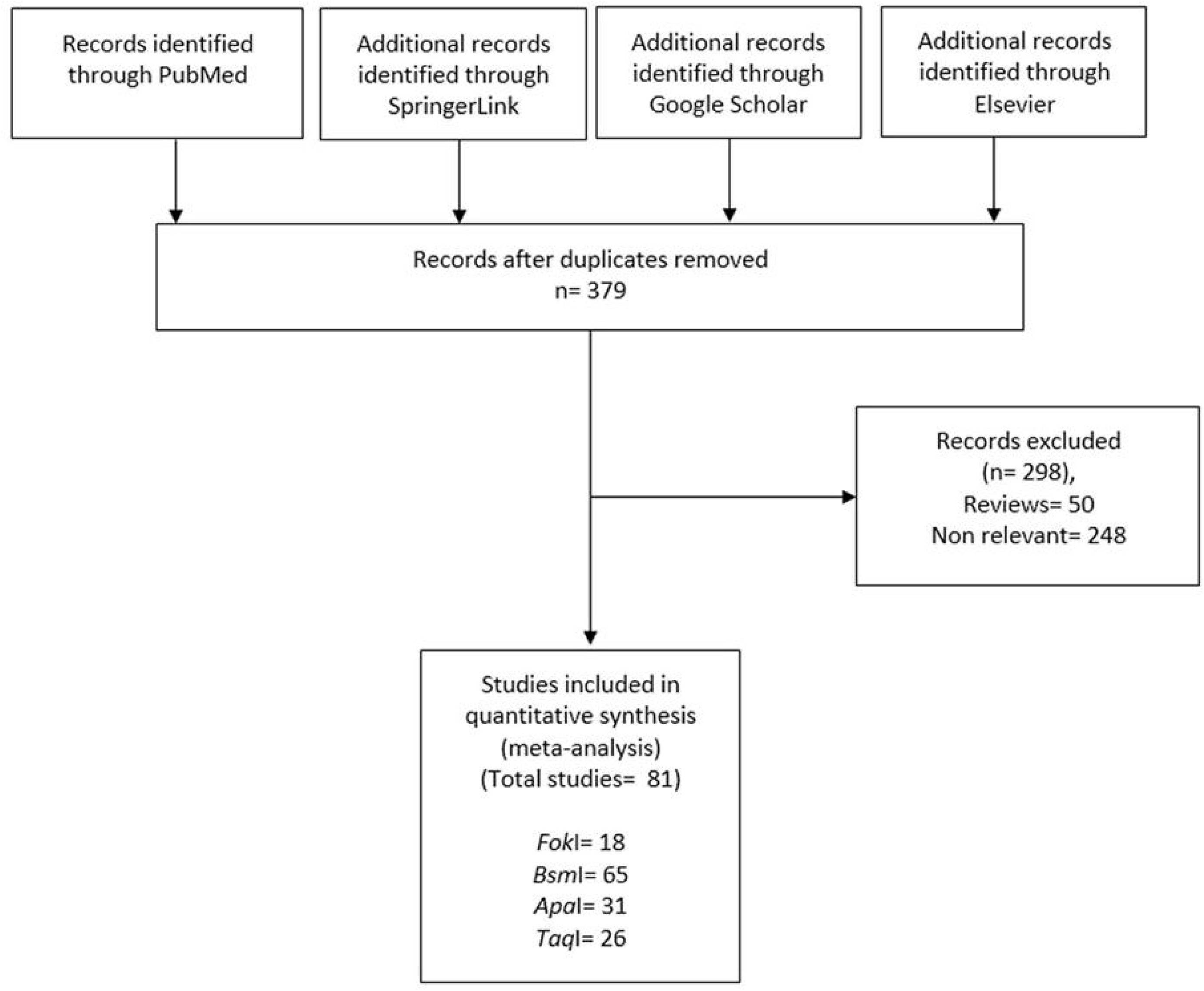
Flow diagram of study search and selection process.

### Eligible studies

For *Bsm*I total 65 studies with 6,880 cases and 8,049 controls were found eligible to include in the meta-analysis [17-81].

For *Apa*I total 31 studies with 3,763 cases and 3,934 controls were found to be eligible to include in the meta-analysis [19, 23, 25, 33, 39, 40, 43, 46, 51, 58, 59, 61, 64, 66, 68, 70, 72, 74, 76, 78-80, 82-90].

For *Fok*I meta-analysis total 18 studies with 1,895 cases and 1,722 controls were found eligible to include in the meta-analysis [33, 40, 45, 51, 56, 62, 65, 66, 68, 70, 74, 76, 79, 91-95].

For *Taq*I total 26 studies including 2,458 cases and 2,895 controls were found eligible to include in the meta-analysis [19, 23, 25, 33, 40, 43, 46, 51, 58, 59, 64, 66, 68, 70, 72, 74, 76, 78-81, 87, 88, 90, 96, 97].

### Meta-analysis

#### *BsmI* meta-analysis

Meta-analysis with allele contrast model showed insignificant association with high heterogeneity (OR_bvsB_= 0.89; 95% CI: 0.78-1.01; p= 0.09; I^2^= 82.02%; P_heterogeneity_= <0.001). No significant association was found in any other genetic model. In dominant model (bb+Bb vs. BB): OR= 0.81, 95% CI= 0.68-0.97, p= 0.02; for homozygote model (bb vs. BB): OR= 0.77, 95% CI= 0.60-0.99, p= 0.04; for co-dominant model (Bb vs. BB): OR= 0.85, 95% CI= 0.73-0.98, p= 0.03; and for recessive model (BB+Bb vs. bb): OR= 0.88, 95% CI= 0.74-1.06, p= 0.20. High heterogeneity was found in all the genetic contrast models except co-dominant model (Table 1; Figure 2).

**Table 1.** Summary estimates for the odds ratio (OR) of *Bsm*I in various allele/genotype contrasts, the significance level (p value) of heterogeneity test (Q test), and the I^2^ metric.

| Gene | Genetic Contrast | Fixed effect<br>OR (95% CI), p | Random effect<br>OR (95% CI), p | Heterogeneity<br>p-value (Q<br>test) | $I^2$ (%) | Publication<br>Bias (p of<br>Egger's test) |
| --- | --- | --- | --- | --- | --- | --- |
| Overall<br>(65) | Allele Contrast (b vs. B) | 0.90 (0.85-0.94), <0.001 | 0.89 (0.78-1.01), 0.09 | <0.001 | 82.02 | 0.73 |
|  | Dominant (bb+Bb vs. BB) | 0.84 (0.77-0.92), <0.001 | 0.81 (0.68-0.97), 0.02 | <0.001 | 65.61 | 0.34 |
|  | Homozygote (bb vs. BB) | 0.81 (0.73-0.90), <0.001 | 0.77 (0.60-0.99), 0.04 | <0.001 | 76.01 | 0.58 |
|  | Co-dominant (Bb vs. BB) | 0.88 (0.80-0.97), 0.01 | 0.85 (0.73-0.98), 0.03 | <0.001 | 43.51 | 0.33 |
|  | Recessive (BB+Bb vs. bb) | 0.89 (0.83-0.96), 0.004 | 0.88 (0.74-1.06), 0.20 | <0.001 | 77.37 | 0.94 |
| Asian<br>(22) | Allele Contrast (b vs. B) | 0.84 (0.74-0.95), 0.008 | 0.86 (0.61-1.19), 0.36 | <0.001 | 81.58 | 0.92 |
|  | Dominant (bb+Bb vs. BB) | 0.70 (0.55-0.90), 0.005 | 0.70 (0.46-1.06), 0.09 | 0.007 | 47.66 | 0.91 |
|  | Homozygote (bb vs. BB) | 0.63 (0.47-0.84), 0.002 | 0.64 (0.34-1.22), 0.17 | <0.001 | 68.37 | 0.70 |
|  | Co-dominant (Bb vs. BB) | 0.77 (0.59-1.00), 0.05 | 0.75 (0.58-0.98), 0.03 | 0.84 | 0 | 0.79 |
|  | Recessive (BB+Bb vs. bb) | 0.86 (0.72-1.03), 0.10 | 0.84 (0.56-1.27), 0.42 | < 0.001 | 75.82 | 0.78 |
| Caucasian<br>(37) | Allele Contrast (b vs. B) | 0.87 (0.82-0.92), <0.001 | 0.86 (0.74-1.00), 0.05 | <0.001 | 81.09 | 0.74 |
|  | Dominant (bb+Bb vs. BB) | 0.85 (0.77-0.94), 0.003 | 0.84 (0.69-1.04), 0.11 | <0.001 | 69.26 | 0.57 |
|  | Homozygote (bb vs. BB) | 0.78 (0.69-0.88), <0.001 | 0.76 (0.57-1.02), 0.06 | <0.001 | 77.36 | 0.63 |
|  | Co-dominant (Bb vs. BB) | 0.91 (0.82-1.02), 0.11 | 0.90 (0.75-1.08), 0.29 | <0.001 | 52.43 | 0.72 |
|  | Recessive (BB+Bb vs. bb) | 0.82 (0.75-0.90), <0.001 | 0.81 (0.66-1.00), 0.05 | <0.001 | 75.5 | 0.72 |
| Other<br>(6) | Allele Contrast (b vs. B) | 1.28 (1.08-1.51), 0.003 | 1.19 (0.76-1.85), 0.43 | <0.001 | 84.7 | 0.45 |
|  | Dominant (bb+Bb vs. BB) | 1.00 (0.75-1.33), 0.96 | 0.82 (0.40-1.67), 0.59 | <0.001 | 80.11 | 0.31 |
|  | Homozygote (bb vs. BB) | 1.50 (1.08-2.10), 0.01 | 1.27 (0.54-3.00), 0.57 | <0.001 | 80.65 | 0.54 |
|  | Co-dominant (Bb vs. BB) | 0.77 (0.57-1.05), 0.10 | 0.62 (0.31-1.24), 0.18 | <0.001 | 75.66 | 0.17 |
|  | Recessive (BB+Bb vs. bb) | 1.69 (1.32-2.16), <0.001 | 1.71 (0.97-3.03), 0.06 | < 0.001 | 78.25 | 0.79 |

**Figure 2.**
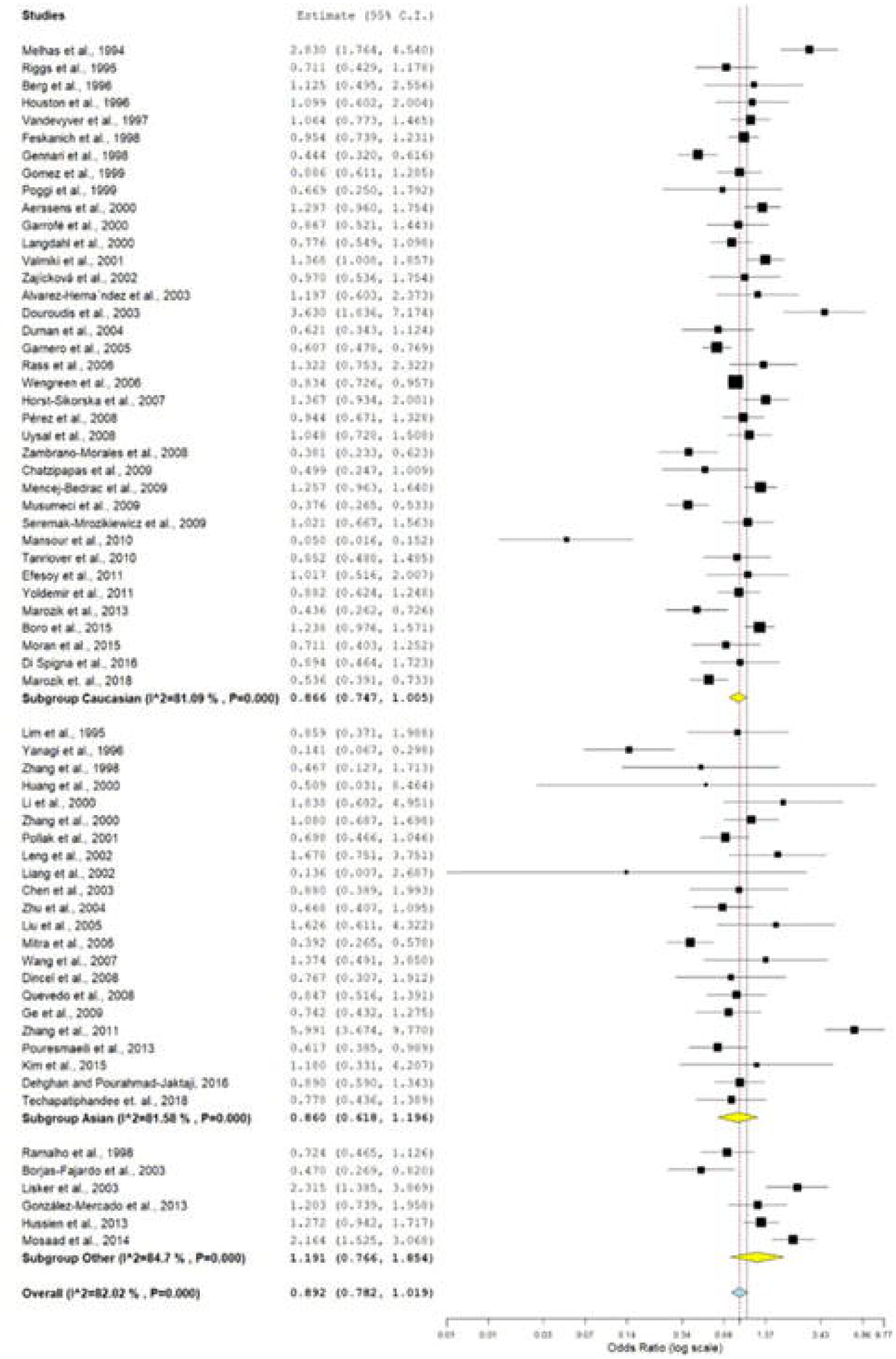
Random effect forest plot of allele contrast model (b vs. B) of VDR *Bsm*I polymorphism. Results of individual and summary OR estimates, and 95% CI of each study were shown. Horizontal lines represented 95% CI, and dotted vertical lines represent the value of the summary OR.

The sub-group analyses were conducted on the basis of ethnicity. Out of 65 studies 37 were belongs to Caucasians, 22 were Asian and six were of other origin. High heterogeneity was observed in almost all genetic models in all sub-groups. No significant association were found in any sub-group analyses in any genetic model (Table 1; Figure 2).

#### *ApaI* meta-analysis

Meta-analysis with allele contrast model showed insignificant association with high heterogeneity (OR_avsA_= 1.01; 95% CI: 0.87-1.17; p= 0.86; I^2^= 74.82%; P_heterogeneity_= <0.001). No significant association was found in any other genetic model. In dominant model (aa+Aa vs. AA): OR= 0.95, 95% CI= 0.78-1.14, p= 0.60; for homozygote model (aa vs. AA): OR= 0.97, 95% CI= 0.72-1.30, p= 0.84; for co-dominant model (Aa vs. AA): OR= 0.92, 95% CI= 0.81-1.04, p= 0.21; and for recessive model (AA+Aa vs. aa): OR= 1.02, 95% CI= 0.81-1.28, p= 0.83 (Table 2; Figure 3).

**Table 2.** Summary estimates for the odds ratio (OR) of *Apa*I in various allele/genotype contrasts, the significance level (p value) of heterogeneity test (Q test), and the I^2^ metric.

| Gene | Genetic Contrast | Fixed effect<br>OR (95% CI), p | Random effect<br>OR (95% CI), p | Heterogeneity<br>p-value (Q<br>test) | $I^2$ (%) | Publication<br>Bias (p of<br>Egger's test) |
| --- | --- | --- | --- | --- | --- | --- |
| Overall<br>(31) | Allele Contrast (a vs. A) | 0.99 (0.92-1.06), 0.90 | 1.01 (0.87-1.17), 0.86 | <0.001 | 74.82 | 0.79 |
|  | Dominant (aa+Aa vs. AA) | 0.92 (0.82-1.04), 0.20 | 0.95 (0.78-1.14), 0.60 | <0.001 | 55.28 | 0.17 |
|  | Homozygote (aa vs. AA) | 0.96 (0.83-1.11), 0.60 | 0.97 (0.72-1.30), 0.84 | <0.001 | 68.58 | 0.65 |
|  | Co-dominant (Aa vs. AA) | 0.92 (0.81-1.04), 0.21 | 0.93 (0.79-1.09), 0.40 | 0.051 | 31.3 | 0.09 |
|  | Recessive (AA+Aa vs. aa) | 1.06 (0.94-1.18), 0.30 | 1.02 (0.81-1.28), 0.83 | <0.001 | 68.95 | 0.50 |
| Asian<br>(12) | Allele Contrast (a vs. A) | 0.99 (0.89-1.12), 0.99 | 1.10 (0.84-1.45), 0.46 | <0.001 | 78.48 | 0.32 |
|  | Dominant (aa+Aa vs. AA) | 1.01 (0.82-1.24), 0.90 | 1.09 (0.81-1.49), 0.54 | 0.05 | 43.75 | 0.04 |
|  | Homozygote (aa vs. AA) | 0.90 (0.71-1.15), 0.43 | 1.03 (0.64-1.65), 0.89 | 0.004 | 60.16 | 0.32 |
|  | Co-dominant (Aa vs. AA) | 1.14 (0.91-1.44), 0.24 | 1.12 (0.89-1.41), 0.32 | 0.83 | 0 | 0.007 |
|  | Recessive (AA+Aa vs. aa) | 0.99 (0.83-1.17), 0.90 | 1.01 (0.70-1.46), 0.93 | <0.001 | 72.92 | 0.82 |
| Caucasian<br>(15) | Allele Contrast (a vs. A) | 0.96 (0.86-1.06), 0.45 | 0.92 (0.72-1.18), 0.54 | <0.001 | 78.00 | 0.49 |
|  | Dominant (aa+Aa vs. AA) | 0.91 (0.77-1.08), 0.31 | 0.90 (0.66-1.23), 0.52 | <0.001 | 67.83 | 0.81 |
|  | Homozygote (aa vs. AA) | 0.94 (0.76-1.17), 0.62 | 0.86 (0.52-1.42), 0.57 | <0.001 | 75.96 | 0.57 |
|  | Co-dominant (Aa vs. AA) | 0.91 (0.77-1.09), 0.34 | 0.91 (0.70-1.19), 0.52 | 0.014 | 49.93 | 0.96 |
|  | Recessive (AA+Aa vs. aa) | 0.98 (0.81-1.18), 0.87 | 0.88 (0.61-1.28), 0.53 | <0.001 | 68.33 | 0.42 |
| Other<br>(4) | Allele Contrast (a vs. A) | 1.07 (0.91-1.26), 0.38 | 1.07 (0.90-1.27), 0.39 | 0.36 | 5.32 | 0.76 |
|  | Dominant (aa+Aa vs. AA) | 0.82 (0.63-1.07), 0.15 | 0.82 (0.62-1.07), 0.15 | 0.43 | 0 | 0.48 |
|  | Homozygote (aa vs. AA) | 1.11 (0.79-1.55), 0.52 | 1.17 (0.64-1.13), 0.60 | 0.03 | 65.55 | 0.44 |
|  | Co-dominant (Aa vs. AA) | 0.67 (0.50-0.89), 0.007 | 0.67 (0.50-0.89), 0.007 | 0.63 | 0 | 0.68 |
|  | Recessive (AA+Aa vs. aa) | 1.42 (1.10-1.83), 0.007 | 1.49 (1.00-2.23), 0.04 | 0.09 | 52.4 | 0.38 |

**Figure 3.**
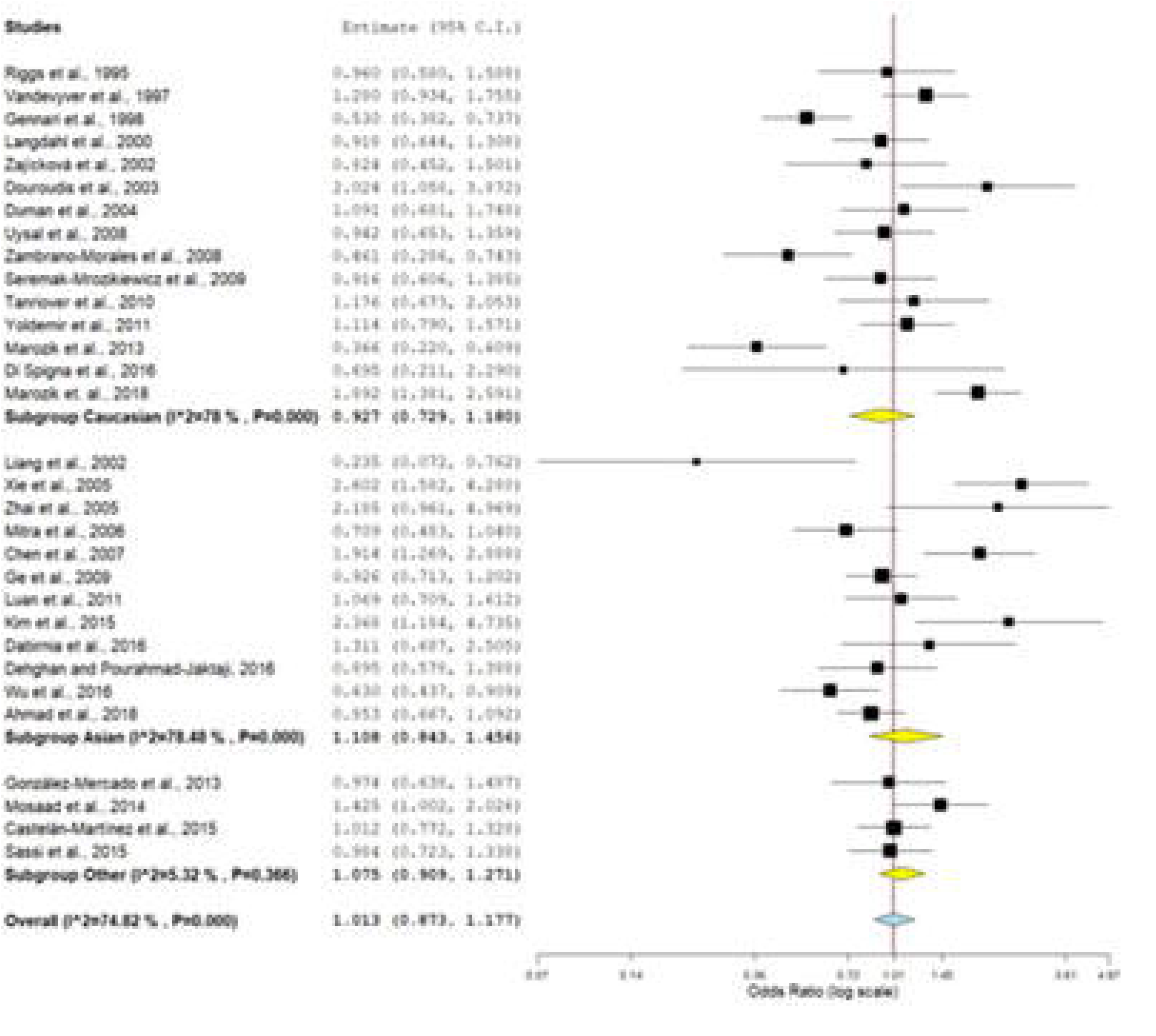
Random effect forest plot of allele contrast model (a vs. A) of VDR *Apa*I polymorphism.

The sub-group analyses were conducted on the basis of ethnicity. Out of 31 studies 15 were belongs to Caucasians, 12 were Asian and four were of other origin. High heterogeneity was observed in Caucasian studies while low heterogeneity was found in Asian and other studies. No significant results were found in any sub-group in any genetic models except for recessive model (AA+Aa vs. aa): OR= 1.49, 95% CI= 1.00-2.23, p= 0.04 in other studies (Table 2; Figure 3).

#### *FokI* meta-analysis

Significant association was found using dominant model (OR_ff+Ffvs.FF_= 1.19, 95% CI: 1.04-1.36, p= 0.01; I^2^= 39.36%). No significant association was observed in any other genetic model (allele contrast model: OR_fvsF_= 1.13,95% CI: 0.95-1.34, p= 0.15, I^2^= 61.8%, P_heterogeneity_= <0.001; homozygote model (ff vs. FF): OR= 1.38, 95% CI= 0.92-2.05, p= 0.11; co-dominant model (Ff vs. FF): OR= 1.12, 95% CI= 0.97-1.30, p= 0.11; and recessive model (FF+Ff vs. ff): OR= 1.34, 95% CI= 0.94-1.91, p= 0.10 (Table 3; Figure 4).

**Table 3.** Summary estimates for the odds ratio (OR) of *Fok*I in various allele/genotype contrasts, the significance level (p value) of heterogeneity test (Q test), and the I^2^ metric.

| *Gene* | *Genetic Contrast* | **Fixed effect**<br>**OR (95% CI), p** | **Random effect**<br>**OR (95% CI), p** | **Heterogeneity**<br>**p-value (Q**<br>**test)** | **I<sup>2</sup>**<br>**(%)** | **Publication Bias**<br>**(p of Egger's test)** |
| --- | --- | --- | --- | --- | --- | --- |
| **Overall**<br>**(18)** | Allele Contrast (f vs. F) | 1.19 (1.08-1.31), <0.001 | 1.13 (0.95-1.34), 0.15 | <0.001 | 61.8 | 0.64 |
|  | Dominant (ff+Ff vs. FF) | 1.19 (1.04-1.36), 0.01 | 1.13 (0.94-1.37), 0.18 | 0.04 | 39.36 | 0.40 |
|  | Homozygote (ff vs. FF) | 1.47 (1.19-1.83), <0.001 | 1.38 (0.92-2.05), 0.11 | <0.001 | 62.08 | 0.99 |
|  | Co-dominant (Ff vs. FF) | 1.12 (0.97-1.30), 0.11 | 1.10 (0.93-1.29), 0.24 | 0.29 | 13.69 | 0.15 |
|  | Recessive (FF+Ff vs. ff) | 1.40 (1.15-1.72), <0.001 | 1.34 (0.94-1.91), 0.10 | 0.001 | 57.98 | 0.69 |
| **Asian**<br>**(5)** | Allele Contrast (f vs. F) | 1.28 (1.07-1.53), 0.007 | 1.17 (0.76-1.82), 0.45 | <0.001 | 79.79 | 0.50 |
|  | Dominant (ff+Ff vs. FF) | 1.24 (0.96-1.59), 0.08 | 1.16 (0.71-1.89), 0.53 | 0.02 | 65.88 | 0.61 |
|  | Homozygote (ff vs. FF) | 1.73 (1.18-2.52), 0.004 | 1.68 (0.68-4.14), 0.25 | <0.001 | 78.92 | 0.88 |
|  | Co-dominant (Ff vs. FF) | 1.15 (0.87-1.52), 0.30 | 1.09 (0.70-1.70), 0.69 | 0.12 | 45.24 | 0.36 |
|  | Recessive (FF+Ff vs. ff) | 1.66 (1.16-2.37), 0.005 | 1.60 (0.76-3.37), 0.21 | 0.004 | 73.74 | 0.88 |
| **Caucasian**<br>**(10)** | Allele Contrast (f vs. F) | 1.31 (0.99-1.29), 0.06 | 1.05 (0.86-1.29), 0.61 | 0.04 | 48.84 | 0.78 |
|  | Dominant (ff+Ff vs. FF) | 1.15 (0.96-1.38), 0.12 | 1.07 (0.83-1.39), 0.57 | 0.09 | 39.58 | 0.65 |
|  | Homozygote (ff vs. FF) | 1.27 (0.95-1.70), 0.10 | 1.11 (0.73-1.70), 0.61 | 0.09 | 39.35 | 0.28 |
|  | Co-dominant (Ff vs. FF) | 1.11 (0.91-1.34), 0.27 | 1.07 (0.85-1.35), 0.54 | 0.22 | 23.34 | 0.63 |
|  | Recessive (FF+Ff vs. ff) | 1.21 (0.92-1.59), 0.15 | 1.12 (0.78-1.61), 0.53 | 0.17 | 29.92 | 0.44 |
| **Other**<br>**(3)** | Allele Contrast (f vs. F) | 1.26 (0.97-1.64), 0.08 | 1.31 (0.84-2.04), 0.21 | 0.07 | 60.97 | 0.58 |
|  | Dominant (ff+Ff vs. FF) | 1.24 (0.86-1.77), 0.23 | 1.24 (0.86-1.77), 0.23 | 0.62 | 0 | 0.82 |
|  | Homozygote (ff vs. FF) | 1.91 (1.00-3.65), 0.05 | 3.28 (0.51-20.87), 0.20 | 0.01 | 78.17 | 0.07 |
|  | Co-dominant (Ff vs. FF) | 1.11 (0.76-1.61), 0.56 | 1.11 (0.76-1.61), 0.56 | 0.74 | 0 | 0.07 |
|  | Recessive (FF+Ff vs. ff) | 1.72 (0.96-3.05), 0.06 | 3.30 (0.49-22.00), 0.21 | 0.005 | 81.08 | 0.001 |

**Figure 4.**
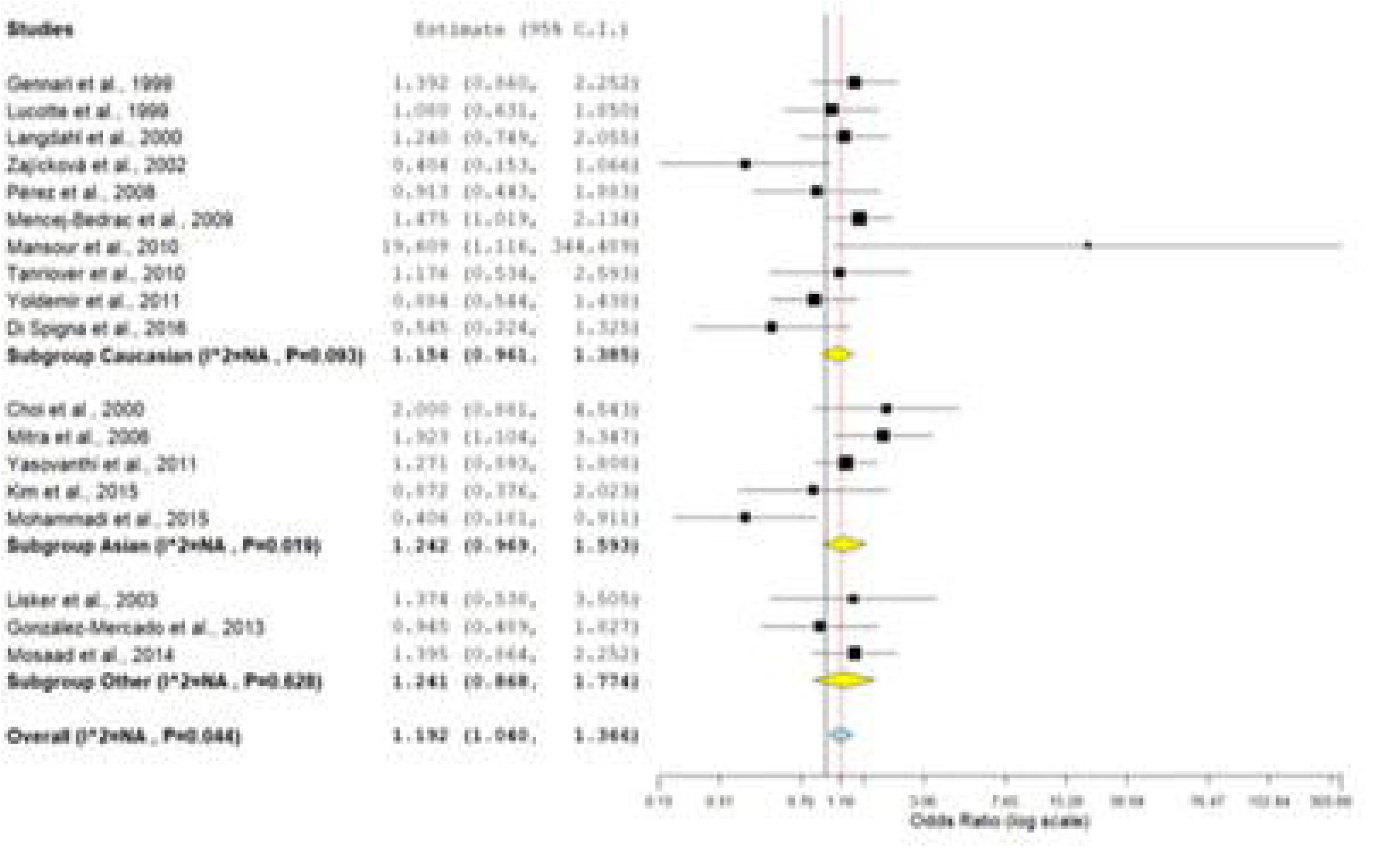
Fixed effect forest plot of dominant model (ff+Ff vs. FF) of VDR *Fok*I polymorphism.

The sub-group analysis was conducted on the basis of ethnicity. Out of 18 studies ten were belongs to Caucasians, five were Asian and three were of other ethnicity. Low heterogeneity was observed in Caucasian studies but high heterogeneity was found in Asian and other studies. No significant results were found in any sub-group in any genetic model (Table 3; Figure 4).

#### *TaqI* meta-analysis

Meta-analysis with allele contrast model showed insignificant association with high heterogeneity (OR_tvsT_= 1.10; 95% CI: 0.91-1.32; p= 0.30; I^2^= 77.26%; P_heterogeneity_= <0.001). High heterogeneity was found in all other genetic models so random effect model was applied. No significant association was found in any other genetic model. In dominant model (tt+Tt vs. TT): OR= 1.09, 95% CI= 0.84-1.41, p= 0.48; for homozygote model (tt vs. TT): OR= 1.20, 95% CI= 0.85-1.69, p= 0.29; for co-dominant model (Tt vs. TT): OR= 1.04, 95% CI= 0.82-1.33, p= 0.70; and for recessive model (TT+Tt vs. tt): OR= 1.16, 95% CI= 0.91-1.48, p= 0.20 (Table 4; Figure 5).

**Table 4.**
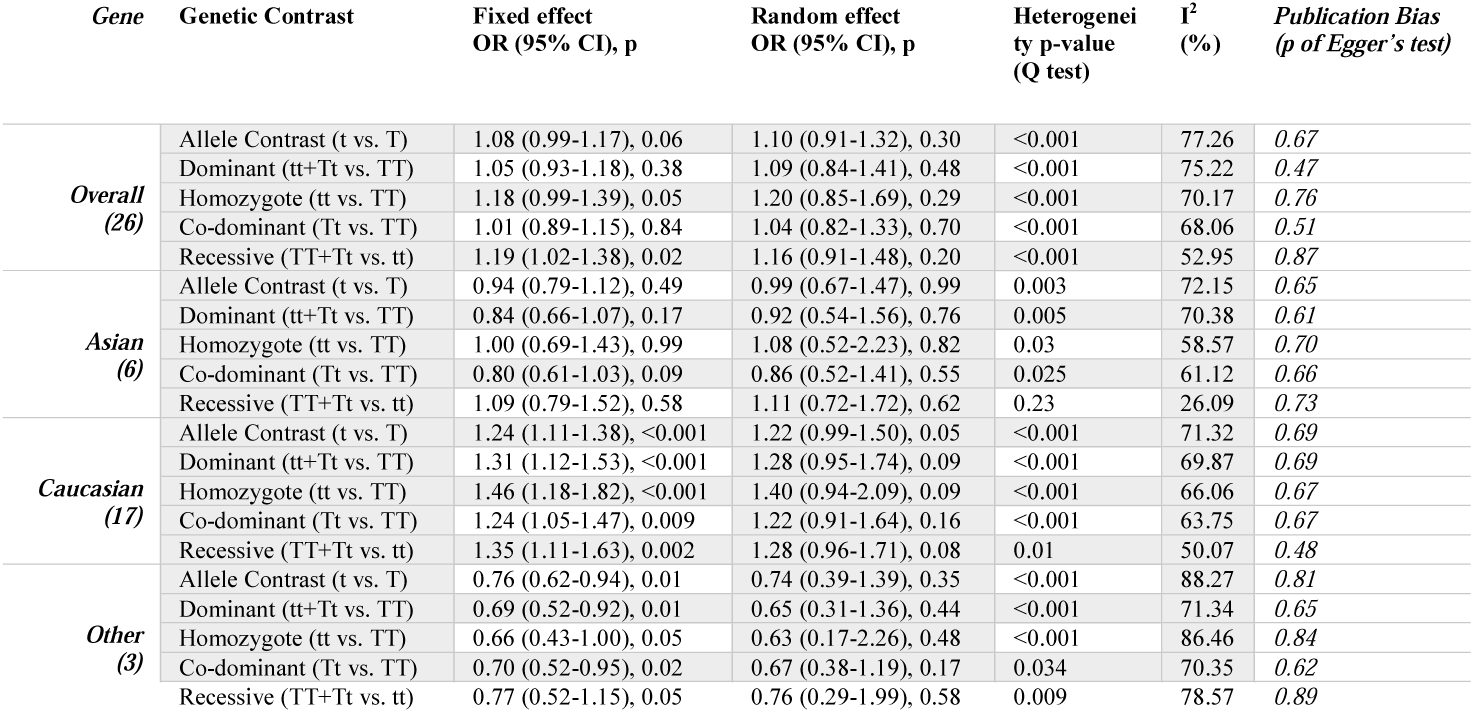
Summary estimates for the odds ratio (OR) of *Taq*I in various allele/genotype contrasts, the significance level (p value) of heterogeneity test (Q test), and the I^2^ metric.

**Figure 5.**
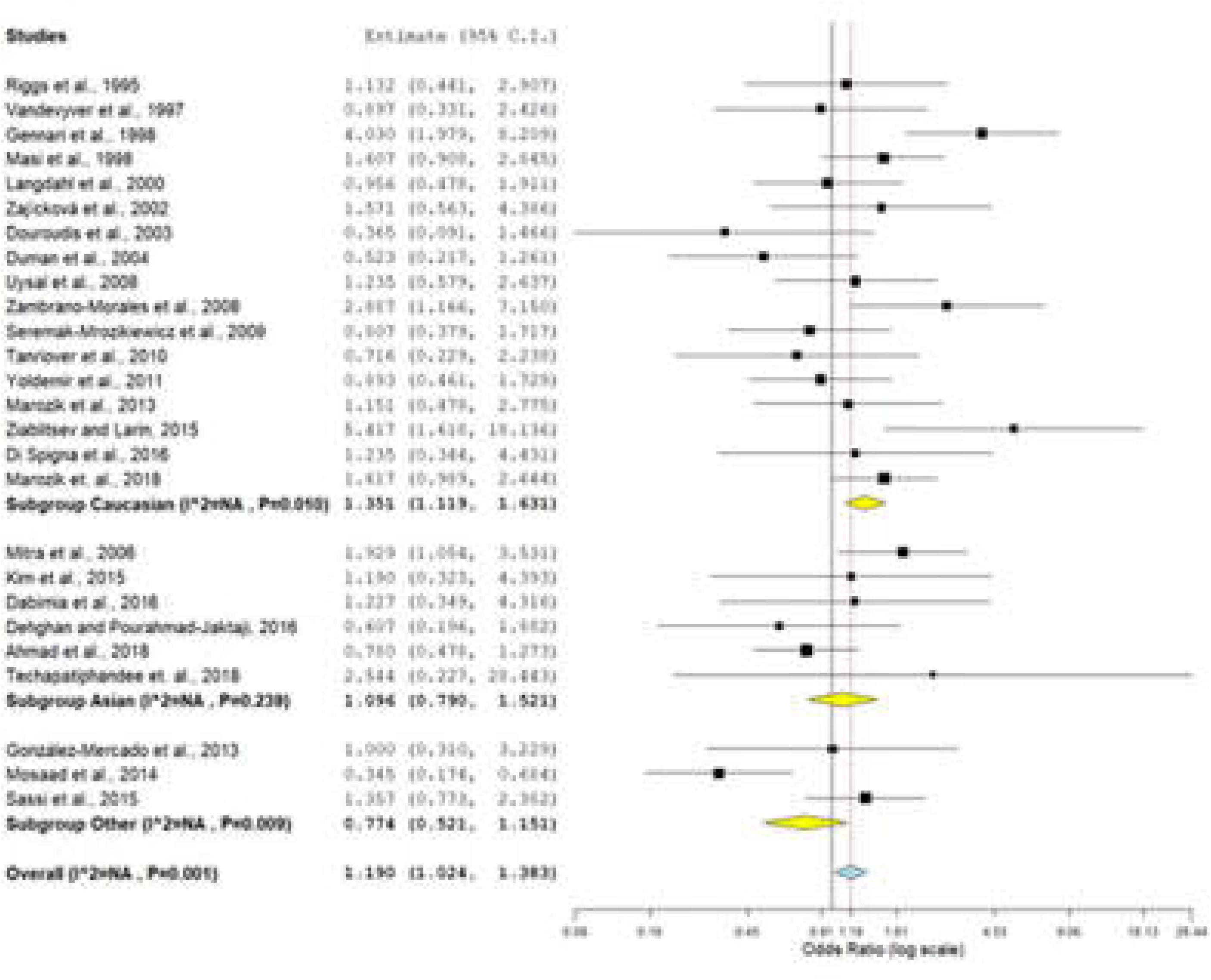
Fixed effect forest plot of recessive model (TT+Tt vs. tt) of VDR *Taq*I polymorphism.

The sub-group analyses were conducted on the basis of ethnicity. Out of 26 studies 17 were belongs to Caucasians, six were Asian and three were of other ethnicity. High heterogeneity was observed in all groups i.e. Asian, Caucasian and other studies. No significant results were found in any sub-group in any genetic models except for recessive model (TT+Tt vs. tt): OR= 1.35, 95% CI= 1.11-1.63, p= 0.002 in Caucasian population (Table 4; Figure 5).

### Sensitivity analysis

Sensitivity analysis was performed by eliminating studies in which control population was not in Hardy Weinberg Equilibrium (p <0.05). The control samples of 21 *Bsm*I studies [22, 25, 29, 33, 34, 39, 43-47, 53, 55, 57, 59, 61, 63, 65, 66, 71, 75] were deviated from the HWE. Sensitivity analysis was performed after removal of these 21 studies and results showed no significant association with osteoporosis risk in the main analysis (OR_bvsB_= 0.99; 95% CI: 0.85–1.15; p= 0.92; I^2^= 77.48%) or in any sub-groups- (Asian subgroup-OR_bvsB_= 0.99; 95% CI: 0.66–1.50; p= 0.99; I^2^= 83.65%: Caucasian subgroup-OR_bvsB_=0.96; 95% CI: 0.83–1.11; p= 0.65; I^2^= 69.61%: other studies subgroup-OR_bvsB_= 1.24; 95% CI: 0.64–2.43; p= 0.51; I^2^= 86.53%). Heterogeneity decreases both in the overall and sub-group meta-analyses except the Asian studies.

In total 18 *Fok*I studies, control population in five studies [51, 65, 74, 94, 95] were not in HWE. After removal of these studies and no significant association was found in the main analysis (OR_fvsF_ = 1.12; 95% CI: 0.99–1.26; p= 0.05; I^2^= 46.48%) or in any sub-groups. Heterogeneity was decreased both in the overall and sub-group meta-analyses.

The control samples of nine *Apa*I studies [23, 25, 39, 43, 46, 51, 66, 78, 89] were not in HWE. Result of meta-analysis after removal of these nine studies showed no association between *Apa*I polymorphism and osteoporosis risk in the main/overall analysis (OR_avsA_= 1.07; 95% CI: 0.90–1.27; p= 0.39; I^2^= 73.94%) and Caucasian population (OR_avsA_=0.85; 95% CI: 0.63–1.16; p= 0.32; I^2^= 78.62%) but the Asian population (OR_avsA_= 1.42; 95% CI: 1.03–1.96; p= 0.03; I^2^= 77.61%); and subgroup other studies (recessive model: OR_AA+Aavs.aa_= 1.49; 95% CI: 1.00–2.23; p= 0.04; I^2^= 52.4%) showed statistically significant association with osteoporosis. Heterogeneity was also decreased both in the overall and sub-group meta-analyses.

Out of 26 *Taq*I studies, control samples of four studies [23, 51, 72, 96] were deviated from the HWE. Results of meta-analysis of 22 studies (after elimination of 4 studies deviated from HWE) did not show any association between Taq1 polymorphism and osteoporosis risk either in total studies (OR_tvsT_= 1.05; 95% CI: 0.85–1.29; p= 0.63; I^2^= 78.86%) or in any sub-group. Moreover, removal of these 4 studies, heterogeneity was increased both in the overall and sub-group meta-analyses except the Asian population.

### Publication bias

The funnel plots are symmetrical for all genetic models in overall and sub-group meta-analyses for all polymorphisms except recessive model of subgroup other studies in *Fok*I and co-dominant model of Asian studies in *Apa*I polymorphisms (Figure 6; Table 1-4). Moreover, Egger’s test reveals no evidence of publication bias in any genetic model in overall meta-analyses of all four polymorphisms.

**Figure 6.**
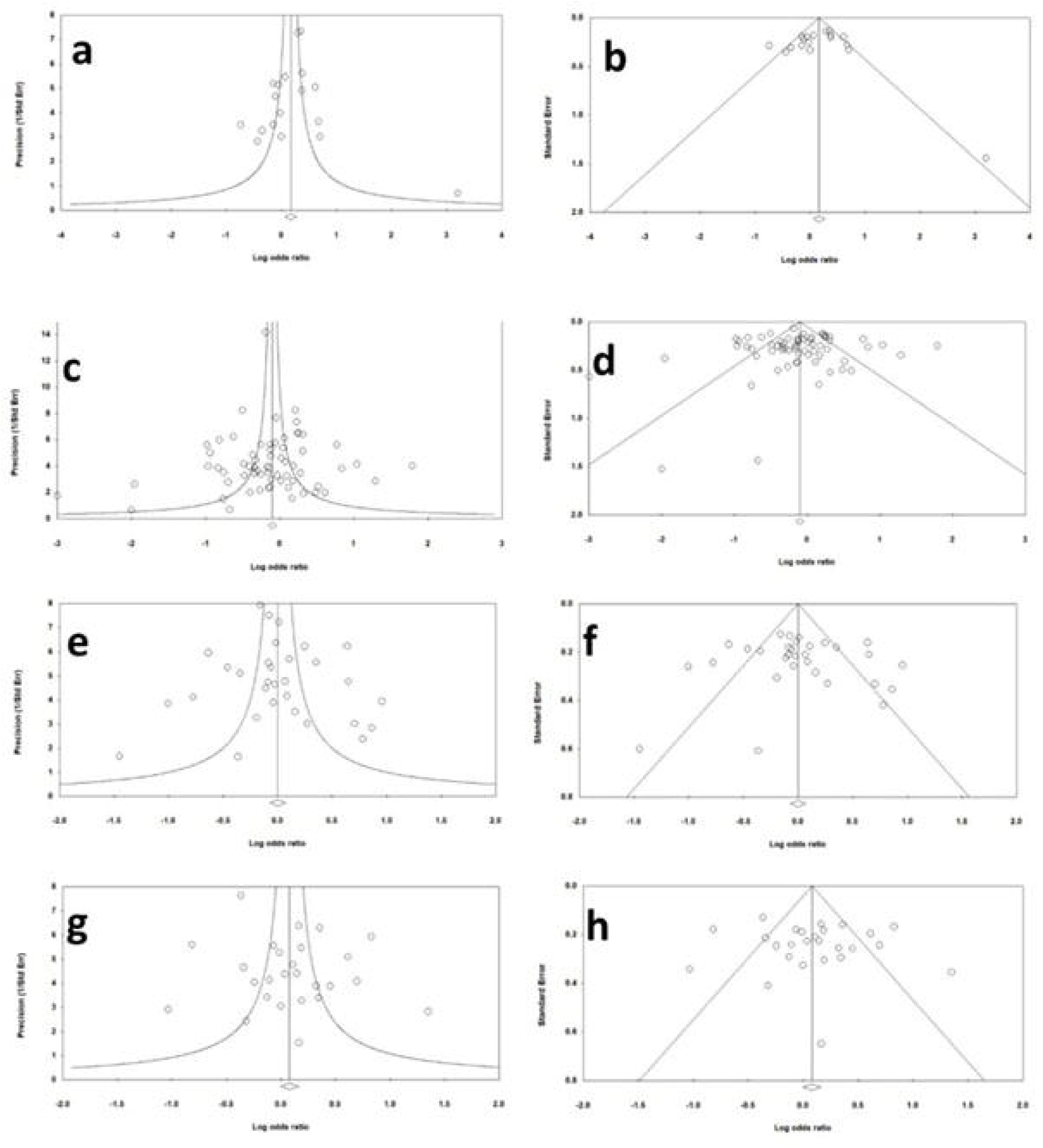
Funnel plots- for *Fok*I a) Precision by log odds ratio, b) standard error by log odds ratio; for *Bsm*I c) Precision by log odds ratio, d) standard error by log odds ratio; for *Apa*I e) Precision by log odds ratio, f) standard error by log odds ratio; for *Taq*I g) Precision by log odds ratio, h) standard error by log odds ratio.

## Discussion

VDRs are members of the nuclear hormone receptor (NR1I) family, which includes pregnane X (PXR) and constitutive androstane receptors (CAR), which form heterodimers with members of the retinoid X receptor family and expressed in the intestine, thyroid and kidney [98]. Vitamin D receptor is considered to be the primary mediator of vitamin D endocrine actions, which can regulate calcium homeostasis and reduce the risk of osteoporosis. VDR translocated from cytoplasm to nucleus upon activation by binding of its ligand 1-α-25-dihydroxyvitamin D3 (1-α-25(OH) 2D3) [99]. Numerous investigations have reported that *VDR* gene polymorphisms were connected with the onset of osteoporosis [81] and other diseases like-breast cancer [100], diabetes [101], myocardial infarction [102] and metabolic syndrome and inflammation [103].

Meta-analysis is a well established statistical tool used for combining the data of small sample sized individual studies. Meta-analysis increases the power of study and decreases type I and II errors. During past two decades, a number of meta-analyses were published which assessed the polymorphism of small effect genes as risk factor for different diseases and disorders e.g. Down syndrome [16], neural tube defects [104], Glucose 6-phosphate dehydrogenase deficiency [105], depression [106], schizophrenia [107], Alzheimer [108], breast cancer [109], colorectal cancer [110], esophageal cancer [111] and prostate cancer [112] etc.

During literature search we identified seven meta-analyses [15, 113-118] investigating the relationship *VDR* gene polymorphism and osteoporosis. *Bsm*I, *Apa*I, *Fok*I and *Taq*I polymorphism were included in seven, four, two and two meta-analyses respectively. *Bsm*I polymorphism studies were included in all seven meta-analyses. Six meta-analyses [15, 113-117] did not report any significant association between osteoporosis susceptibility and *Bsm*I polymorphism. Zhang et al [118] conducted a meta-analysis on the risk of osteoporosis in postmenopausal women with 36 studies including 7,192 subjects and found marginally significant association (OR_bvs.B_= 1.2; CI= 1.00-1.46; p= 0.052). In all the meta-analyses high between studies heterogeneity was not found except the study conducted by Yu et al [115]. *Apa*I polymorphism was included in four meta-analyses [113, 115, 117, 118]. Zintzaras et al [113], Yu et al [115], Wang et al [117] and Zhang et al [118] included seven, six, three and 18 studies respectively in their meta-analyses and all four studies reported no association between *Apa*I polymorphism and osteoporosis risk. Zintzaras et al [113] and Zhang et al [118] conducted meta-analyses of three and 18 studies of *Fok*I polymorphism and they did not found significant association between *Fok*I polymorphism and osteoporosis. Both groups [113, 118] also conducted meta-analyses of *Taq*I polymorphism studies and again reported no association between *Taq*I polymorphism and osteoporosis susceptibility.

In the present meta-analysis, four common VDR gene polymorphisms (*Bsm*I, *Apa*I, *Fok*I and *Taq*I), largest sample size and highest number of studies were included. total 65 (14,929 samples), 31 (7,697 samples), 18 (3,617 samples) and 26 (5,353 samples) studies for *Bsm*I, *Apa*I, *Fok*I and *Taq*I polymorphisms respectively were included. We found significant association dominant model of *Fok*I polymorphism (ff+Ff vs. FF: OR= 1.19, 95% CI= 1.04-1.36, p= 0.01) with low heterogeneity (I^2^= 39.36). No association was found in sub-group analysis on the basis of ethnicity in any genetic model except in the Caucasian population in the recessive model of *Taq*I polymorphism (TT+Tt vs. tt: OR= 1.35, 95% CI= 1.11-1.63, p= 0.002) with moderate heterogeneity (I^2^= 50.07).

The present meta-analysis has few demerits like- (i) used crude odds ratio, (ii) only genetic polymorphisms considered, other factors such as environmental factors or food habits are not included which might have important role in the etiology of osteoporosis. With these limitations the present study has strength also like- (i) this is largest meta-analysis conducted both in number of included studies and number of sample size (81 studies; 19,268 samples), (ii) included all common VDR polymorphisms (*Bsm*I, *Apa*I, *Fok*I and *Taq*I).

## Conclusion

In conclusion, we found the *Fok*I polymorphism is associated with osteoporosis also the *Taq*I polymorphism is a risk factor for the Caucasian population. While other polymorphisms (*Bsm*I and *Apa*I) has no role in the etiology of osteoporosis in total or stratified populations. Furthermore, we suggest that for the future case-control studies gene–gene and gene–environment interactions should also be considered which might well elucidate genetics of osteoporosis.

## Data Availability

All the data are provided in the manuscript.

## Data Availability

All the data are provided in the manuscript.

## Acknowledgments

Upendra Yadav is highly grateful to VBS Purvanchal University, Jaunpur for providing financial assistance to him in the form of PDF.

## Ethical Approval

The article does not contain any studies with human or animal subjects performed by any of the authors.

